# Core outcome set for clinical trials of stent implantation in coronary heart disease -- a systematic review and mixed-method consensus study

**DOI:** 10.1101/2024.07.09.24310182

**Authors:** Xingli Wan, Xinyi Wang, Xia Li, Youlin Long, Zhengchi Li, Liang Du

## Abstract

**Background:** Although various types of stent interventions have been found in clinical trials to be effective in treating coronary heart disease, researchers often use heterogeneous outcome measures. This can preclude the comparison of results across studies and prevent effective meta-analyses from answering important clinical questions. The need to standardize outcome measure reporting is urgent. This study aims to produce a core outcome set(COS) for coronary stent implantation research.

**Methods:** We performed a systematic review via the PubMed and EMBASE databases and then synthesized recommendations from international standardized endpoint definitions and consensus documents to generate a list of potential outcomes. We conducted a two-round Delphi survey, achieving the iterative selection of potential outcomes. A multidisciplinary panel of experts and patients with coronary stent implantation experience rated the importance of candidate outcomes on a 9-point Likert scale. To be provisionally accepted, outcomes needed to be critical important (score 7-9, with 9 being the maximum) by 70% of each stakeholder group. Finally, we held a consensus meeting to discuss and generate the COS.

**Results:** A total of seventy-two potential outcomes were identified by systematic literature review. We obtained nine key consensus documents to inform the standardized definitions of the outcomes. After duplication and combination of similar outcomes, 26 of the 72 outcomes assigned into five outcome domains were included in the Delphi surveys. Twenty outcomes were provisionally accepted. Following the consensus meetings, the agreement was reached on a twelve-item COS.

**Conclusion:** We developed a COS for coronary stent implantation in coronary heart disease. Its implementation will facilitate the standardization reporting of clinical trials and reduce the heterogeneity in assessment in future research. Further research may help determine the specific, validated measurement instruments to assess each outcome.

**What is Known:**

- Coronary stent implantation is consider to be the primary method for treating coronary heart disease.
- Many trials use heterogeneous effectiveness and safety outcome measures with different defined and non-patient concerned outcome measures, which would largely prevent meta-analyses and contribute to selective reporting bias.

**What the Study Adds:**

- We initially develop the first core outcome set(COS) of coronary stent implantation intervention for China to facilitate the standardization reporting of clinical trials and enable meta-analysis.
- In our study, patient concerned outcome measure were thoroughly discussed and included in the COS by extensively involved patient representatives in every step of the project.
- Although we used rigorous method to development the COS, it should be updated as new evidence emerges in this rapidly evolving field.

## 1 INTRODUCTION

Coronary atherosclerotic heart disease(CHD), which commonly results from severely narrow coronary atherosclerotic lesions, is one of the most lethal diseases globally.^[1]^ The global CHD mortality is projected to grow from 7.594 million in 2016 to about 9.245 million in 2030.^[2]^ The mortality rate attributed to CHD among urban residents in China was 121.59 per 100,000 in 2019, accounting for nearly 50% of the deaths from cardiovascular disease.^[3]^ This rate has been on the rise since 2012.^[4]^

Percutaneous coronary intervention (PCI), especially coronary stent implantation, is the primary method for treating coronary heart disease.^[5]^ Remarkably, given that stent treatment has different properties, the clinical trial is required to report adequately on the effectiveness and safety of stent therapy. ^[6]^ The need to standardize outcome measure reporting is crucial. Given the high case fatality in CHD, stent implantation clinical trials require robust outcome measures that capture what matters to patients, researchers, and professionals.

Unfortunately, many trials use heterogeneous effectiveness and safety outcome measures with different defined^[7,8]^ and non-patient concerned outcome measures^[9]^, which would largely prevent meta-analyses. Lack of outcome standardization may also contribute to selective reporting bias, which may skew estimates of treatment effectiveness.^[10]^ However, the International Consortium for Health Outcomes Measurement (www.ICHOM.org) and COMET (Core Outcome Measures in Effectiveness Trials) have no information about appropriate outcome measures for CHD with stent implantation.^[11]^

A core outcome set (COS) describes a standardized set of outcomes that should be measured and reported in all studies in a specific area, at a minimum.^[12]^ COS can standardize outcome selection, collection, and reporting and represent different vital stakeholders’ input and opinions. Significantly, COS does not limit the number of reportable outcomes because, besides the core set, researchers may report additional outcomes of interest.^[13]^

This study aimed to develop a COS for use in clinical trials, systematic reviews, and clinical practice guidelines evaluating coronary stent implantation interventions for patients with CHD.

## 2 Methods

The study methodology was informed by the COMET Handbook,^[12]^ utilizing the Core Outcome Set-STAndards for Development (COS-STAD) recommendations and Core Outcome Set-STAndards for Reporting (COS-STAR) standards.^[14,15]^ In brief, we used a mixed-method consensus approach to develop this COS involving three stages: (1) Generation of a potential core outcomes list through a literature review of published systematic review, including randomized controlled coronary stent implantation trials. Additional outcomes were added to the list following the supplementary search of relevant guidelines or expert consensus related to coronary artery stent implantation. (2) A two-round Delphi process based on the potential core outcomes list. (3) Patient and medical professional consensus meeting to identify the final core outcome set for coronary stent implantation for CHD.

### 2.1 Participants

We selected a Delphi panel to ensure a broad multi-stakeholder representation. Participants were invited nationwide through the professional contacts of the leading investigator. Guided by the established core outcome set methodology,^[16]^ we aimed to recruit at least 30 participants from each key stakeholder group, with critical consideration including experts with a deep understanding of the project. Therefore, in addition to considering stakeholder inclusion, we formulated the criteria for optimal representation. Eligible participants were:

Healthcare professionals: (i) Clinicians who have worked in the cardiovascular medicine/ surgery ward for over ten years. Considering the geographical distribution, we recruited those clinicians widely from China’s east, south, west, north, and central regions. Moreover, the clinicians who have one of the following conditions are thought to be the best representatives: (1) Have participated in at least two stent implantation clinical trials (as an investigator or participated in the trial’s design) or the research experience is more than three years; (2) Serving as department director or manager, and with excellent academic influence in the cardiovascular field. (ii) Nursing staff: more than ten years in the cardiovascular medicine/surgery ward or as a ward manager with rich experience caring for CHD. (iii) Medical technicians who have worked in cardiac ultrasonography or cardiac imaging wards and have over three years of research experience.

COS users: (i) Chief editor or academic editor of a journal of cardiovascular disease. An editor with a medical background is the best representative of this. (ii) Head of the Marketing Department of a coronary stent industry. Those who have worked for more than five years and have medical background knowledge are the best representatives. (iii) An evidence-based medicine methodologist with cardiovascular disease research experience or a methodologist of COS research is needed. The one with both is the best representative.

Patients with CHD: anyone who has undergone successful coronary stent treatment, is in the postoperative follow-up period during the investigation, and has a good educational background, understanding, and communication skills. Those with medical background knowledge, methodological knowledge, or even both, are the best representatives.

### 2.2 Literature Review

To identify potential core outcomes, we conducted a literature review in the PubMed and EMBASE databases for systematic reviews evaluating coronary stent treatment for CHD from inception until December 2021. We excluded the systematic reviews that included non-RCTs and repeat publishment, as well as individual patient data (IPD) analysis, updated meta-analysis, network meta-analysis, meta-regression analysis, and Bayesian meta-analysis. Searches were limited to English-language publications (or those translated into English).

Two evaluators read abstracts and full texts independently using Microsoft Access 2013. If a consensus could not be reached, a third reviewer arbitrated. We assessed the studies meeting the inclusion criteria using a standardized constructed data-extraction file (Microsoft Access 2013), and the following information was collected: year of study, journal, sample size, participant characteristics, type of intervention(s), type of disease, all reported outcomes, preset primary and secondary outcome.

Two evaluators jointly duplicated, standardized, and merged the outcomes extracted from the systematic review to form a list of original outcomes.

Following the supplementary search of relevant guidelines or expert consensus on coronary artery stent implantation in CNKI, WanFang Data, and COMET, we added additional outcomes to the list.

We assigned the candidate outcomes to one of the five core areas according to the outcome domain classification criteria of Core Outcome Measures in Effectiveness Trials (COMET).[11] This framework represents the following five core areas that outcomes should cover to ensure full breadth of reporting: mortality, pathophysiological manifestations, patient report, life impact, and resource utilization. Furthermore, we differed the candidate outcomes regarding effectiveness or safety outcomes by guiding principles for clinical trials of coronary drug-eluting stents.^[17]^ A list of potential core outcomes for coronary stent implantation clinical trials was finally formed.

### 2.3 Consensus Process

#### 2.3.1 Two-round Delphi Survey

We undertook a prospective consensus study using a Delphi process. In round 1, participants rated the importance of each potential core outcome using a nine-point Likert scale under the Grading of Recommendations Assessment Development and Evaluations (GRADE) working group ^[18]^, where a score of 1-3 indicates limited importance, a score of 4-6 indicates the importance and a score of 7-9 indicates critical importance. Outcomes for each area were presented in a non-random order, each outcome accompanied by a lay definition. Participants could also add outcomes that they felt were significant but did not appear in the preliminary list. We reviewed all outcomes, including those that fulfilled the priori-defined consensus criteria into round 2. New outcomes proposed by at least two participants were taken forward to round 2. We also collected participants’ demographic data, including gender, age, years of work and research experience, workplace, educational background, professional title, and job position.

In round 2, participants received their previous score and the responses of each stakeholder group in the previous round, and whereafter they could rescore outcomes. If the participant rescored the outcome from “limited importance” in round 1 to “critical importance” in round 2 or from “critical importance” to “limited importance,” they would provide the reason.

For the stakeholders of healthcare professionals and COS users, we sent the Delphi questionnaire through email, accompanied by audio explanations of the research objectives and instructions for completing the questionnaire. Each round remained open for two weeks, with regular 1-week reminders (through email, text messages, or WeChat) sent to those who had partially completed or not completed the survey to maximize response rates. Regarding the patient group, we conducted a face-to-face survey with the assistance of personnel responsible for the follow-up of CHD patients at a tertiary hospital’s outpatient clinic in Chengdu, Sichuan Province. While explaining outcome definitions for the patients, we particularly emphasized that scoring the importance of outcomes should be based on the extent of impact on themselves.

#### 2.3.2 Consensus Meeting

We invited participants who had completed two rounds of the Delphi survey to contribute to the consensus meeting. Patient representatives were also present. We held the meeting online to maximize attendance during the continuing COVID-19 pandemic. The meeting included a short presentation that outlined the literature review and the Delphi survey findings, including the final list of outcomes by category and how each stakeholder group voted for them. Attendees reviewed and thoroughly discussed the outcomes included in the consensus meeting and did not reach a consensus in the Delphi surveys, then rescored the outcome using a nine-point Likert scale. We collated the vote results. Based on the prior consensus criteria, all outcomes retained were included in the final COS.

### 2.4 Consensus Definition

The consensus was defined as “a prior.”We strived to gather as many potential core outcomes as possible through Delphi Round 1. In round 2, we aimed to achieve the highest possible level of convergence for the outcomes through an iterative screen. Therefore, for the Delphi process, “Consensus in” for round 1 was defined as≥70% of the whole stakeholders scored 7-9 and ≤ 15% scored 1-3 or ≥ 70% of the participants of any stakeholder group scoring 7-9 and≤15% scoring 1-3; “Consensus in” for Delphi round 2 was defined as ≥70% of participants per stakeholder group scored 7-9 and ≤ 15% scored 1-3; “Consensus out” was defined as at least 70% of the whole stakeholders scoring 1-3 and less than 15% scoring 7-9. Like the Delphi survey, “Consensus in” for consensus meeting was defined as ≥70% of participants per stakeholder group scored 7-9 and ≤ 15% scored 1-3 in each group.

## 3 Results

Figure 1 outlines a flow chart of the results for the number of participants and outcomes in the COS development process.

**Figure 1:**
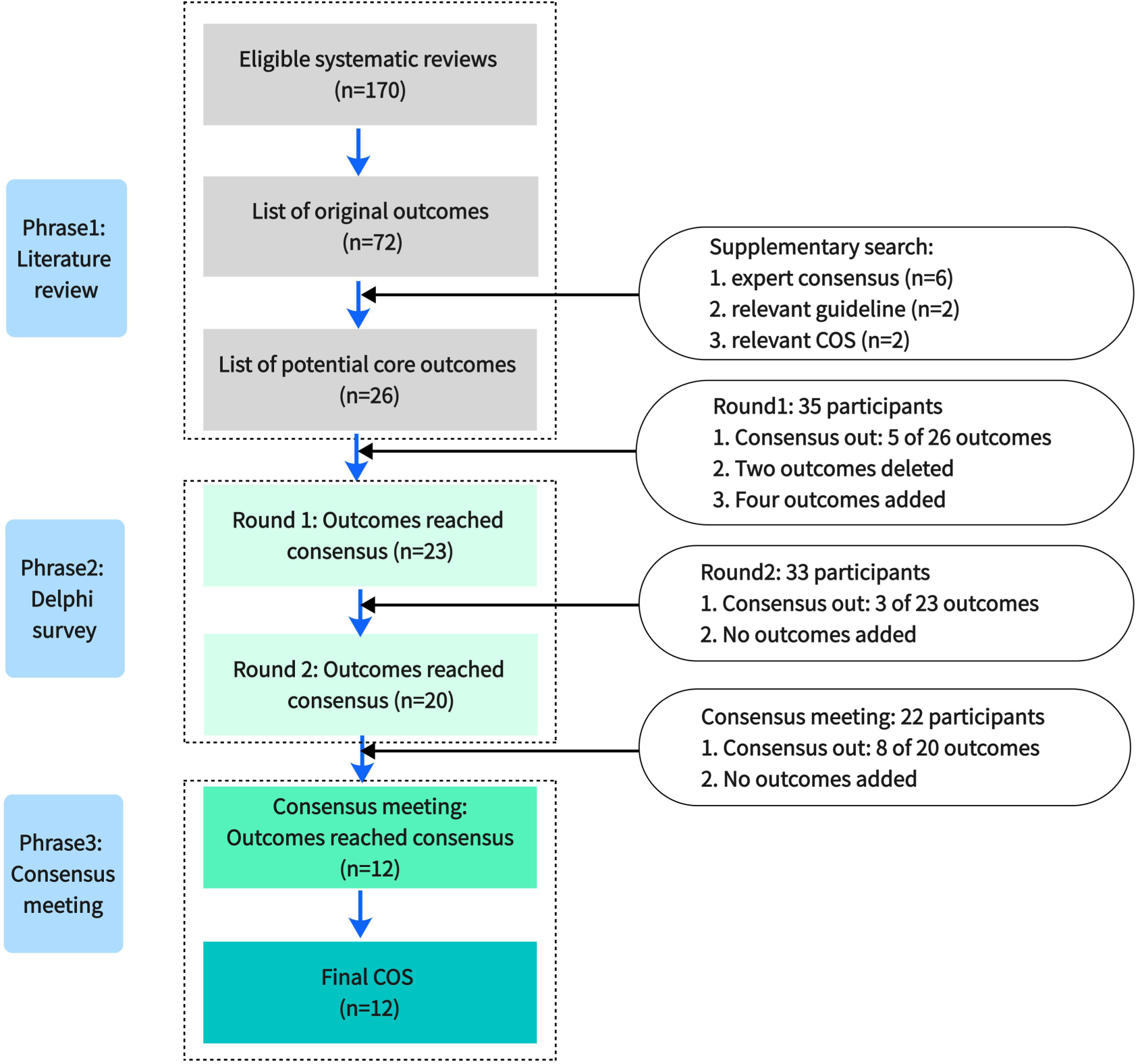
Flow chart of COS development process. COS, core outcome set.

### 3.1 Participants

Thirty-five participants completed the first Delphi survey, followed by a participation rate of 94.3% (*n*=33) in round 2, with seventeen and sixteen optimal representatives included in the first and second Delphi processes, respectively. The thirty-three participants in Roud 2 of Delphi comprised twenty-three experts from two stakeholders (healthcare professionals and COS users) and ten patients. The twenty-three experts from ten nationwide administrative regions covered China’s east, south, west, north, and middle regions. Twenty-three participants attended the consensus meeting, including twelve optimal representatives. Table 1 shows the participation rate and the number of optimal representatives per stakeholder group.

**Table 1.**
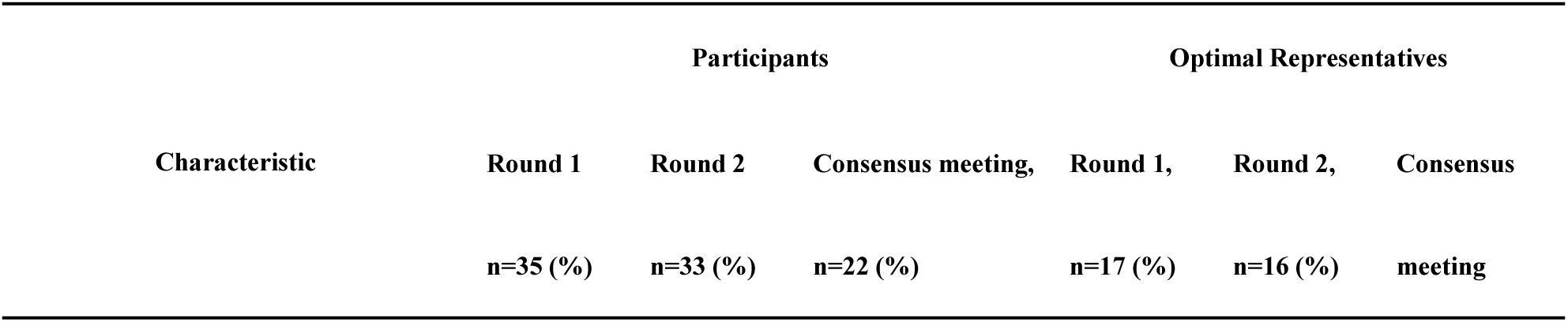
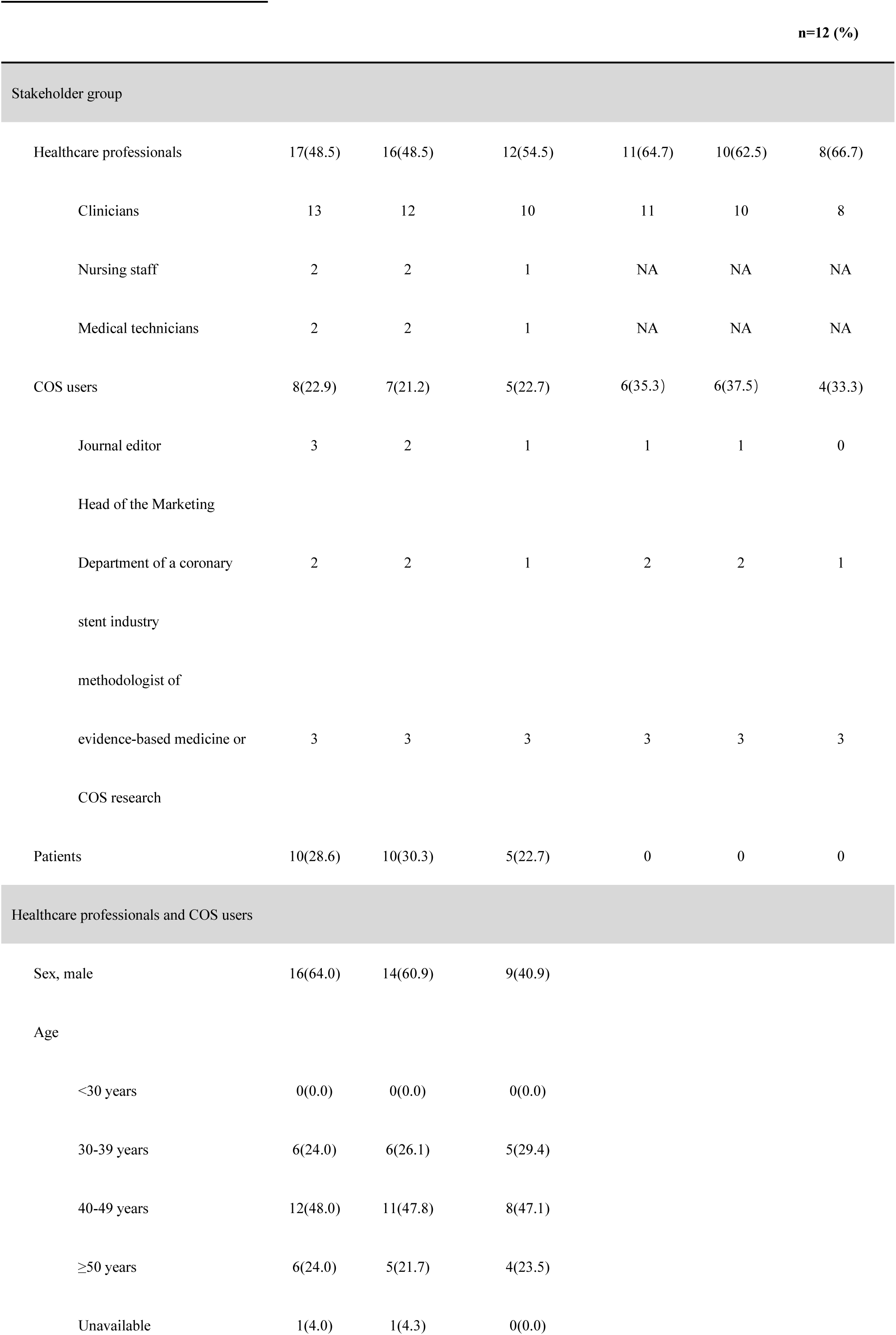
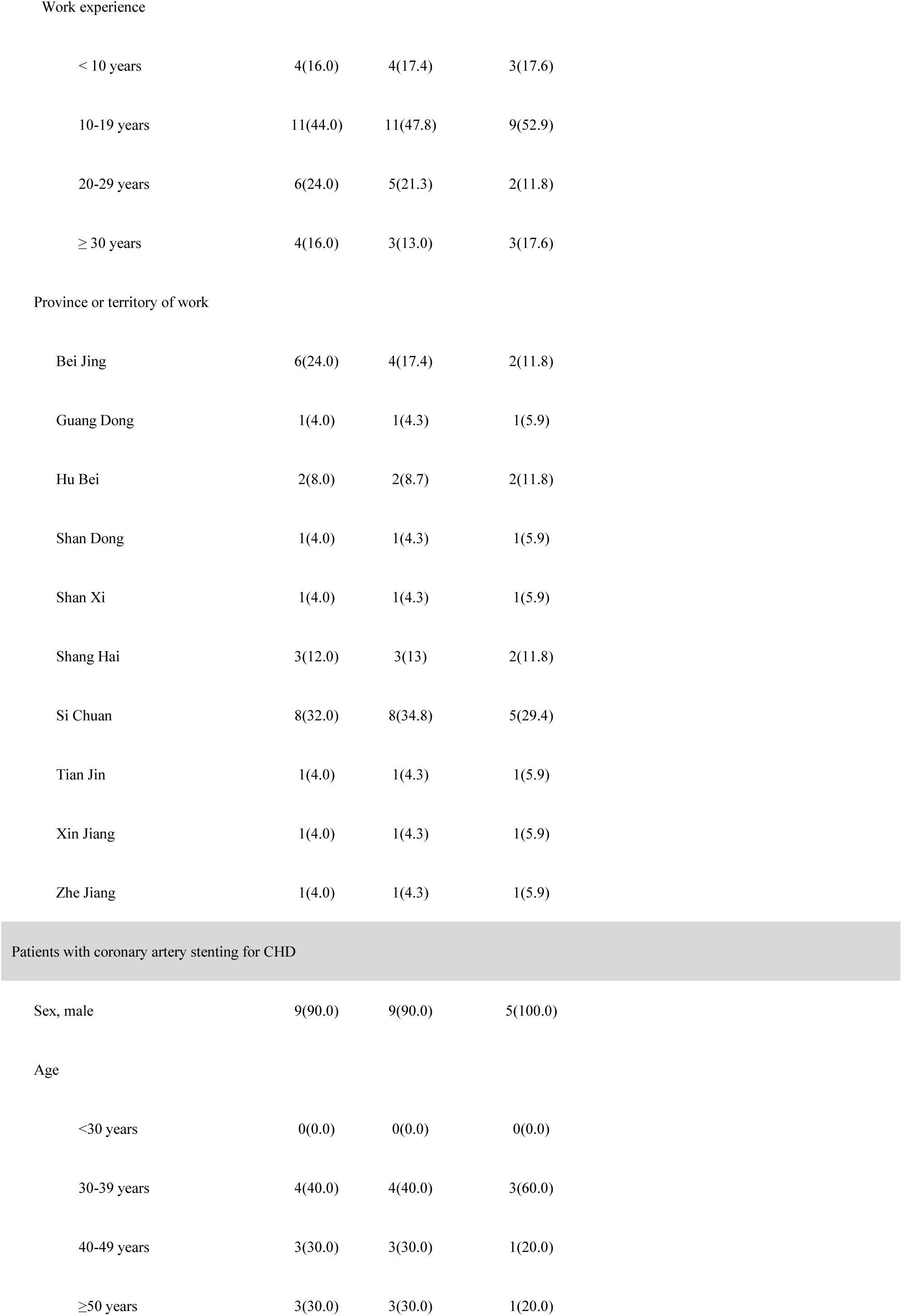
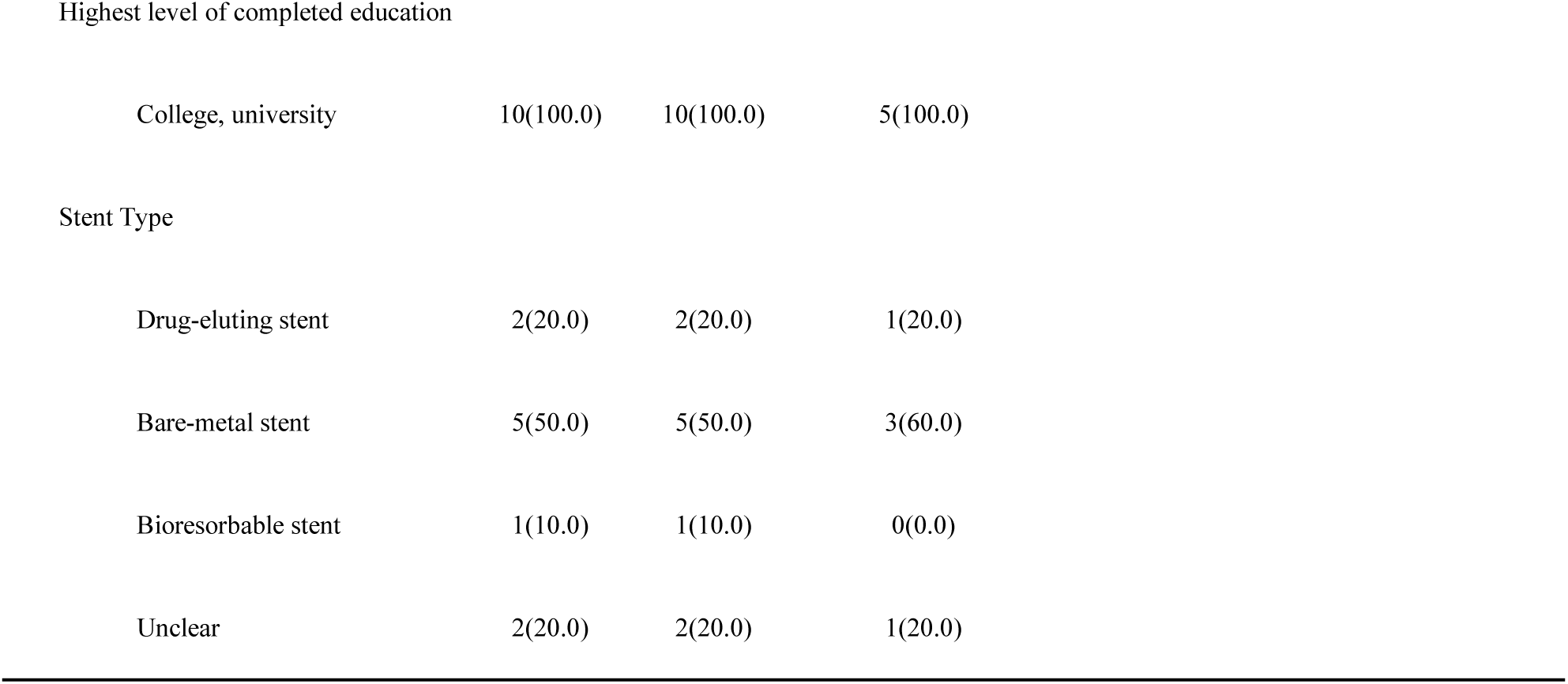
Participation rate and optimal representatives in the Delphi process.

| Characteristic | Participants |  |  | Optimal Representatives |  |  |
| --- | --- | --- | --- | --- | --- | --- |
|  | Round 1 | Round 2 | Consensus meeting, | Round 1, | Round 2, | Consensus |
|  | n=35 (%) | n=33 (%) | n=22 (%) | n=17 (%) | n=16 (%) | meeting |

n=12 (%)
| Stakeholder group |  |  |  |  |  |  |
| --- | --- | --- | --- | --- | --- | --- |
| Healthcare professionals | 17(48.5) | 16(48.5) | 12(54.5) | 11(64.7) | 10(62.5) | 8(66.7) |
| Clinicians | 13 | 12 | 10 | 11 | 10 | 8 |
| Nursing staff | 2 | 2 | 1 | NA | NA | NA |
| Medical technicians | 2 | 2 | 1 | NA | NA | NA |
| COS users | 8(22.9) | 7(21.2) | 5(22.7) | 6(35.3) | 6(37.5) | 4(33.3) |
| Journal editor | 3 | 2 | 1 | 1 | 1 | 0 |
| Head of the Marketing |  |  |  |  |  |  |
| Department of a coronary | 2 | 2 | 1 | 2 | 2 | 1 |
| stent industry |  |  |  |  |  |  |
| methodologist of |  |  |  |  |  |  |
| evidence-based medicine or | 3 | 3 | 3 | 3 | 3 | 3 |
| COS research |  |  |  |  |  |  |
| Patients | 10(28.6) | 10(30.3) | 5(22.7) | 0 | 0 | 0 |
| Healthcare professionals and COS users |  |  |  |  |  |  |
| Sex, male | 16(64.0) | 14(60.9) | 9(40.9) |  |  |  |
| Age |  |  |  |  |  |  |
| <30 years | 0(0.0) | 0(0.0) | 0(0.0) |  |  |  |
| 30-39 years | 6(24.0) | 6(26.1) | 5(29.4) |  |  |  |
| 40-49 years | 12(48.0) | 11(47.8) | 8(47.1) |  |  |  |
| ≥50 years | 6(24.0) | 5(21.7) | 4(23.5) |  |  |  |
| Unavailable | 1(4.0) | 1(4.3) | 0(0.0) |  |  |  |

Work experience
|  |  |  |  |
| --- | --- | --- | --- |
| < 10 years | 4(16.0) | 4(17.4) | 3(17.6) |
| 10-19 years | 11(44.0) | 11(47.8) | 9(52.9) |
| 20-29 years | 6(24.0) | 5(21.3) | 2(11.8) |
| ≥ 30 years | 4(16.0) | 3(13.0) | 3(17.6) |

Province or territory of work
|  |  |  |  |
| --- | --- | --- | --- |
| Bei Jing | 6(24.0) | 4(17.4) | 2(11.8) |
| Guang Dong | 1(4.0) | 1(4.3) | 1(5.9) |
| Hu Bei | 2(8.0) | 2(8.7) | 2(11.8) |
| Shan Dong | 1(4.0) | 1(4.3) | 1(5.9) |
| Shan Xi | 1(4.0) | 1(4.3) | 1(5.9) |
| Shang Hai | 3(12.0) | 3(13) | 2(11.8) |
| Si Chuan | 8(32.0) | 8(34.8) | 5(29.4) |
| Tian Jin | 1(4.0) | 1(4.3) | 1(5.9) |
| Xin Jiang | 1(4.0) | 1(4.3) | 1(5.9) |
| Zhe Jiang | 1(4.0) | 1(4.3) | 1(5.9) |

Patients with coronary artery stenting for CHD
|  |  |  |  |
| --- | --- | --- | --- |
| Sex, male | 9(90.0) | 9(90.0) | 5(100.0) |

Age
|  |  |  |  |
| --- | --- | --- | --- |
| <30 years | 0(0.0) | 0(0.0) | 0(0.0) |
| 30-39 years | 4(40.0) | 4(40.0) | 3(60.0) |
| 40-49 years | 3(30.0) | 3(30.0) | 1(20.0) |
| ≥50 years | 3(30.0) | 3(30.0) | 1(20.0) |

Highest level of completed education
|  |  |  |  |
| --- | --- | --- | --- |
| College, university | 10(100.0) | 10(100.0) | 5(100.0) |

Stent Type
|  |  |  |  |
| --- | --- | --- | --- |
| Drug-eluting stent | 2(20.0) | 2(20.0) | 1(20.0) |
| Bare-metal stent | 5(50.0) | 5(50.0) | 3(60.0) |
| Bioresorbable stent | 1(10.0) | 1(10.0) | 0(0.0) |
| Unclear | 2(20.0) | 2(20.0) | 1(20.0) |

### 3.2 Outcomes

#### 3.2.1 Phase 1: Outcome Identification

We extracted seventy-two outcomes through the literature review, many of which were variations on assessments for coronary stent implantation. We obtain nine important references ^[19-27]^ of international standardized endpoint definitions and consensus documents through supplementary search and extract standardized outcomes added to the list of original outcomes after duplication. This process produced a list of twenty-six potential core outcomes [Tables S1] based on the recommendations of the COMET handbook ^[12]^ that the development of core outcome sets should consider outcomes that matter to patients, which are often endpoint outcomes, i.e., the endpoint outcomes of diseases.

#### 3.2.2 Phase 2: Delphi survey The Delphi process - Round 1

The Delphi survey ran from July to September 2022. Of the twenty-six potential core outcomes, the five highest-scored outcomes in round 1 were all-cause death (mean score 8.8), myocardial infarction (mean score 8.5), TLR (mean score 8.4), TVR (mean score 8.2), and stent thrombosis (mean score 7.9). Twenty-one outcomes achieved the “Consensus in” standard [Table 2].

**Table 2.**
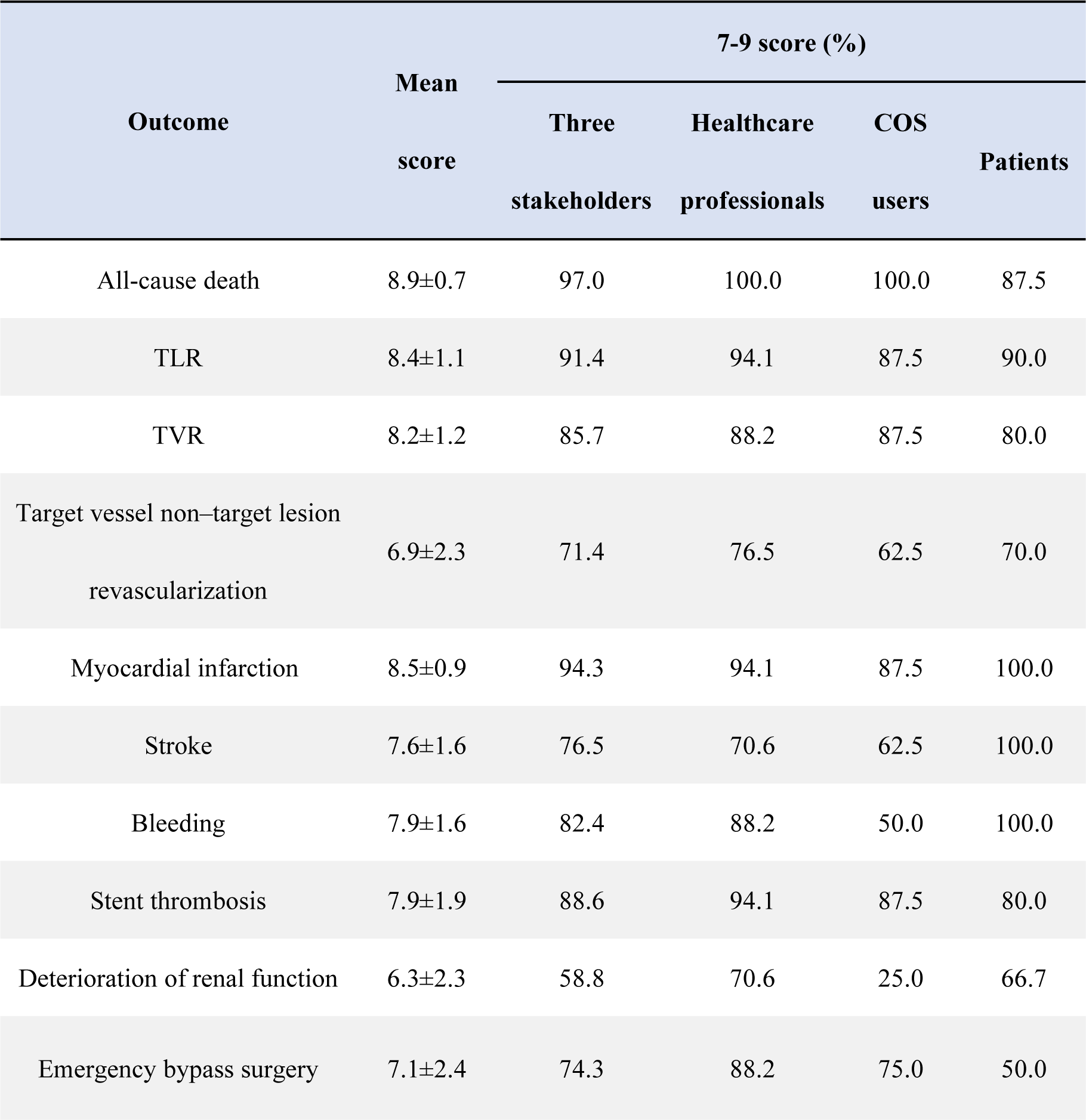
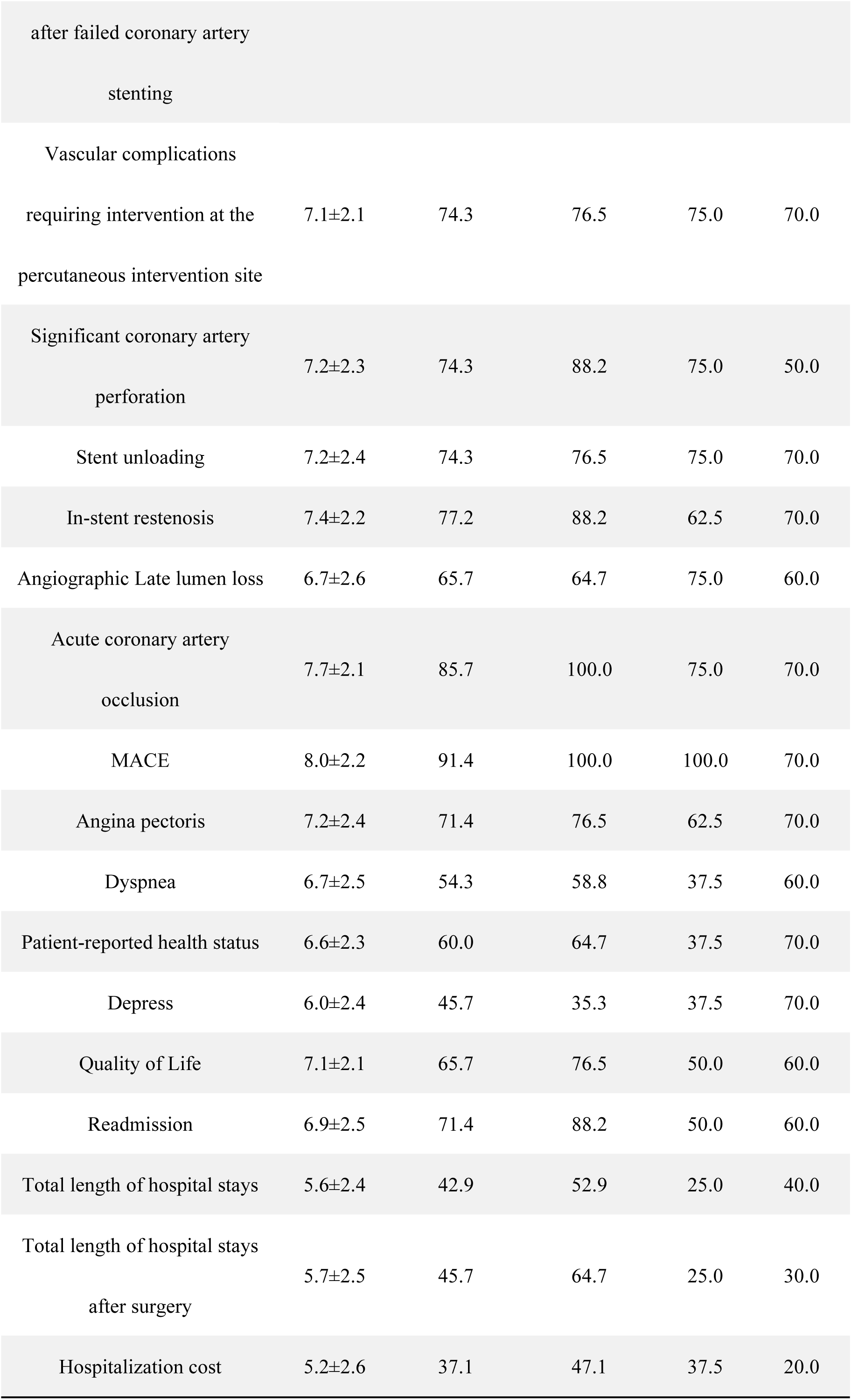
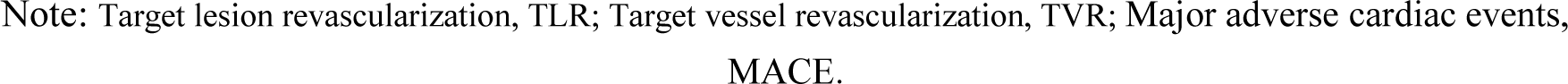
Score results of the participants in Delphi round one.

| Outcome | Mean score | 7-9 score (%) |  |  |  |
| --- | --- | --- | --- | --- | --- |
|  |  | Three stakeholders | Healthcare professionals | COS users | Patients |
| All-cause death | 8.9±0.7 | 97.0 | 100.0 | 100.0 | 87.5 |
| TLR | 8.4±1.1 | 91.4 | 94.1 | 87.5 | 90.0 |
| TVR | 8.2±1.2 | 85.7 | 88.2 | 87.5 | 80.0 |
| Target vessel non–target lesion revascularization | 6.9±2.3 | 71.4 | 76.5 | 62.5 | 70.0 |
| Myocardial infarction | 8.5±0.9 | 94.3 | 94.1 | 87.5 | 100.0 |
| Stroke | 7.6±1.6 | 76.5 | 70.6 | 62.5 | 100.0 |
| Bleeding | 7.9±1.6 | 82.4 | 88.2 | 50.0 | 100.0 |
| Stent thrombosis | 7.9±1.9 | 88.6 | 94.1 | 87.5 | 80.0 |
| Deterioration of renal function | 6.3±2.3 | 58.8 | 70.6 | 25.0 | 66.7 |
| Emergency bypass surgery | 7.1±2.4 | 74.3 | 88.2 | 75.0 | 50.0 |
| after failed coronary artery<br>stenting |  |  |  |  |  |
| Vascular complications<br>requiring intervention at the<br>percutaneous intervention site | 7.1±2.1 | 74.3 | 76.5 | 75.0 | 70.0 |
| Significant coronary artery<br>perforation | 7.2±2.3 | 74.3 | 88.2 | 75.0 | 50.0 |
| Stent unloading | 7.2±2.4 | 74.3 | 76.5 | 75.0 | 70.0 |
| In-stent restenosis | 7.4±2.2 | 77.2 | 88.2 | 62.5 | 70.0 |
| Angiographic Late lumen loss | 6.7±2.6 | 65.7 | 64.7 | 75.0 | 60.0 |
| Acute coronary artery<br>occlusion | 7.7±2.1 | 85.7 | 100.0 | 75.0 | 70.0 |
| MACE | 8.0±2.2 | 91.4 | 100.0 | 100.0 | 70.0 |
| Angina pectoris | 7.2±2.4 | 71.4 | 76.5 | 62.5 | 70.0 |
| Dyspnea | 6.7±2.5 | 54.3 | 58.8 | 37.5 | 60.0 |
| Patient-reported health status | 6.6±2.3 | 60.0 | 64.7 | 37.5 | 70.0 |
| Depress | 6.0±2.4 | 45.7 | 35.3 | 37.5 | 70.0 |
| Quality of Life | 7.1±2.1 | 65.7 | 76.5 | 50.0 | 60.0 |
| Readmission | 6.9±2.5 | 71.4 | 88.2 | 50.0 | 60.0 |
| Total length of hospital stays | 5.6±2.4 | 42.9 | 52.9 | 25.0 | 40.0 |
| Total length of hospital stays<br>after surgery | 5.7±2.5 | 45.7 | 64.7 | 25.0 | 30.0 |
| Hospitalization cost | 5.2±2.6 | 37.1 | 47.1 | 37.5 | 20.0 |

Note: Target lesion revascularization, TLR; Target vessel revascularization, TVR; Major adverse cardiac events, MACE.

Based on the discussion of the selection criteria for core outcome sets, we removed one composite outcome (MACE) and one surrogate endpoint indicator (late lumen loss, LLL) from the twenty-one consensus outcomes. Meanwhile, we consolidated the suggestions proposed by the participants [Table 3]. Four new outcomes (cardiac function, wire-related complications, stent fracture, and cardiovascular death) were added. Finally, twenty-three outcomes were put forward to Delphi round 2.

**Table 3.** Consolidation of the suggestions proposed by participants in Delphi round one.

| Item | Describe |
| --- | --- |
| Consolidation | Merged the two outcomes (coronary artery dissection and coronary artery hematoma) proposed by the participants and the existing outcome of coronary artery injury-related complications as one item. |
|  | Added the outcome of aortic-related complications suggested by participants into the existing outcome, "vascular puncture-related complications". |
| Modification | Reselected the definition of myocardial infarction using a generic definition.<br>The classification of Grade III and above bleeding requiring medical intervention was kept in the definition, removing the Grade I and II classification. |
| Separation | Extracted cardiovascular death from all-cause mortality as a separate outcome. |

##### The Delphi process - Round 2

Of the twenty-three outcomes, the five highest-scored outcomes in round 2 were cardiovascular death (mean score 8.9), all-cause death (mean score 8.8), TLR (mean score 8.5), myocardial infarction (mean score 8.4), and TVR (mean score 8.3). No new outcomes were introduced. Following round 2, three outcomes reached the “Consensus out” criteria, and twenty outcomes that achieved the “Consensus in” standard were eligible to be put forward to the consensus meeting [Figure 2].

**Figure 2:**
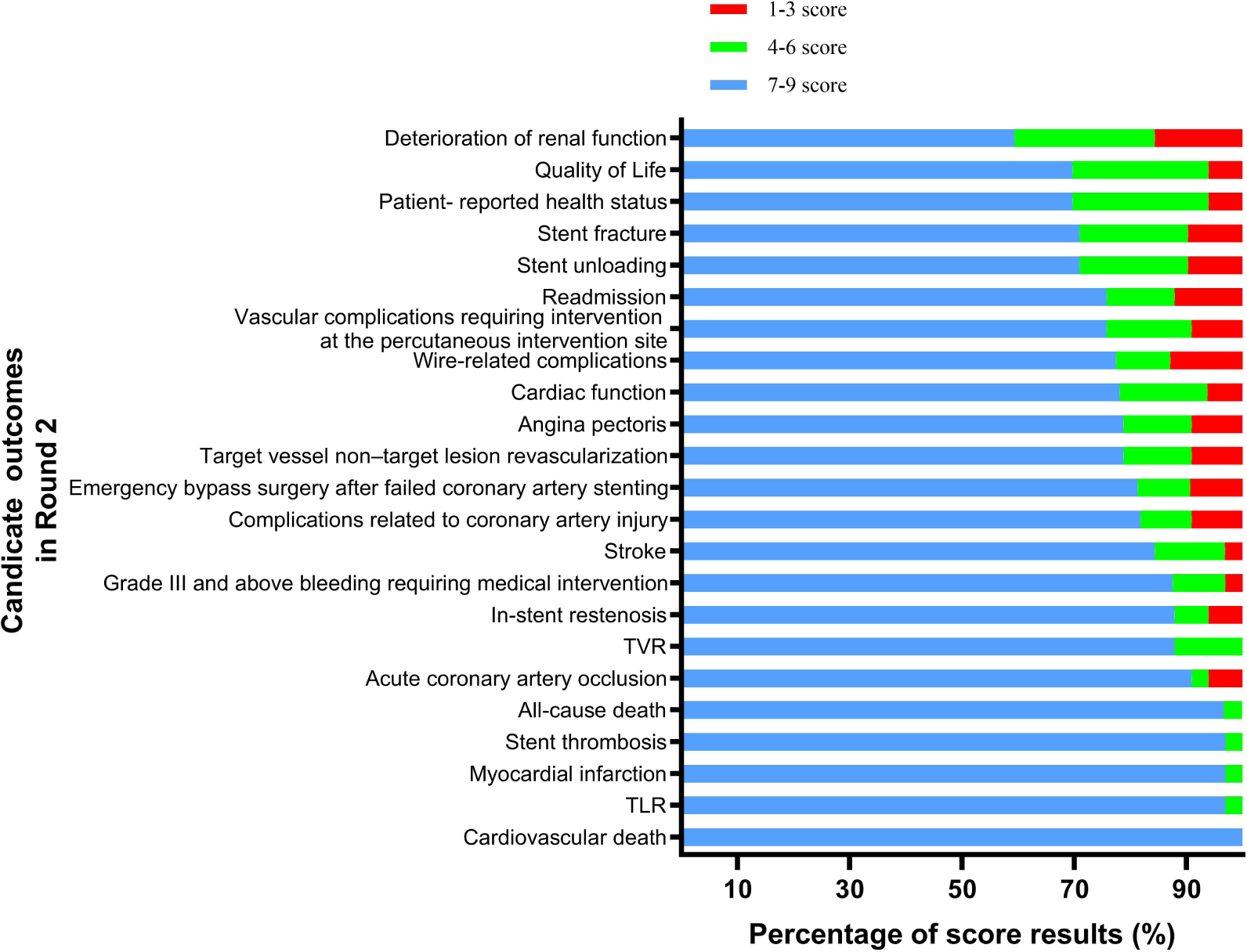
Score results of the candidate outcomes in Delphi round two.

#### 3.2.3 Phase 3: consensus meeting

We held a consensus meeting online on 30 September 2022 because of the COVID-19 pandemic in Chengdu. Twenty-three stakeholders who previously completed the two rounds of the Delphi process attended the meeting. Four outcomes were discussed during the meeting. TLR, TVR, and target vessel/non-target lesion revascularization are outcomes with similar meanings but different levels of importance. TLR is most correlated with the outcomes after stent implantation, while target vessel/non-target lesion revascularization refers to new lesions that require intervention in the target vessels that have received stent implantation, and TVR includes both. The discussion results did not change the previous rankings of the importance of these three outcomes. Stent thrombosis is a crucial outcome of coronary stent implantation, of which severe cases can cause large-scale myocardial infarction in patients. However, bare-metal stents have the highest incidence of stent thrombosis. With the continuous iteration of stents, drug-eluting stents have dramatically reduced the incidence of stent thrombosis. Experts agreed that other more critical outcomes may replace stent thrombosis. Finally, stakeholders rescored these candidate outcomes [Figure 3].

**Figure 3:**
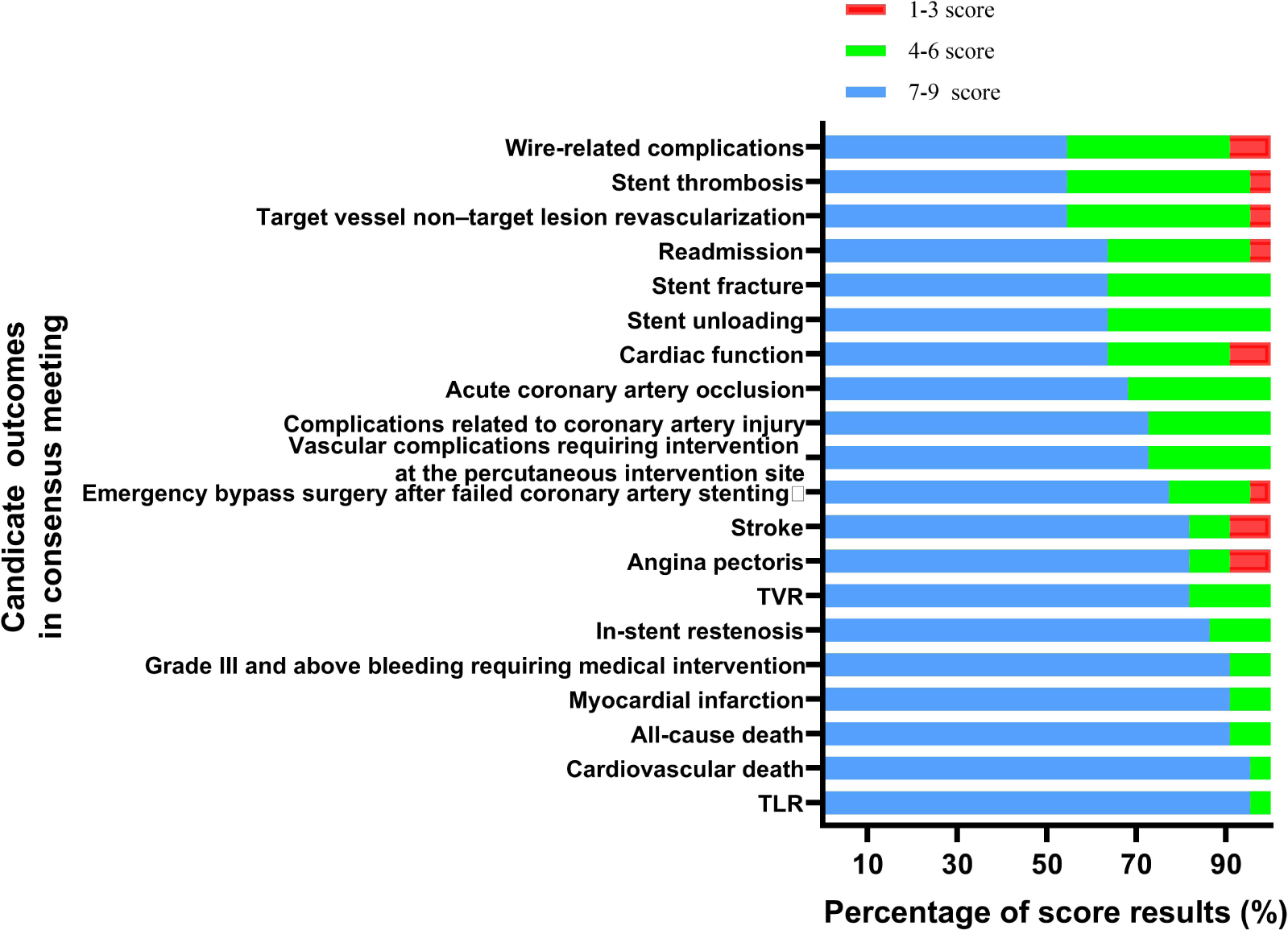
Score results of the candidate outcomes in the consensus meeting.

#### 3.2.3 COS

Twenty-two of the participants voted for the candidate outcomes. The final core outcome set contained twelve outcomes: TLR, cardiovascular death, all-cause death, myocardial infarction, Grade III and above bleeding requiring medical intervention, in-stent restenosis, TVR, angina pectoris, stroke, emergency bypass surgery after failed coronary artery stenting, vascular complications requiring intervention at the percutaneous intervention site, and complications related to coronary artery damage [Figure 4 and Tables S2].

**Figure 4:**
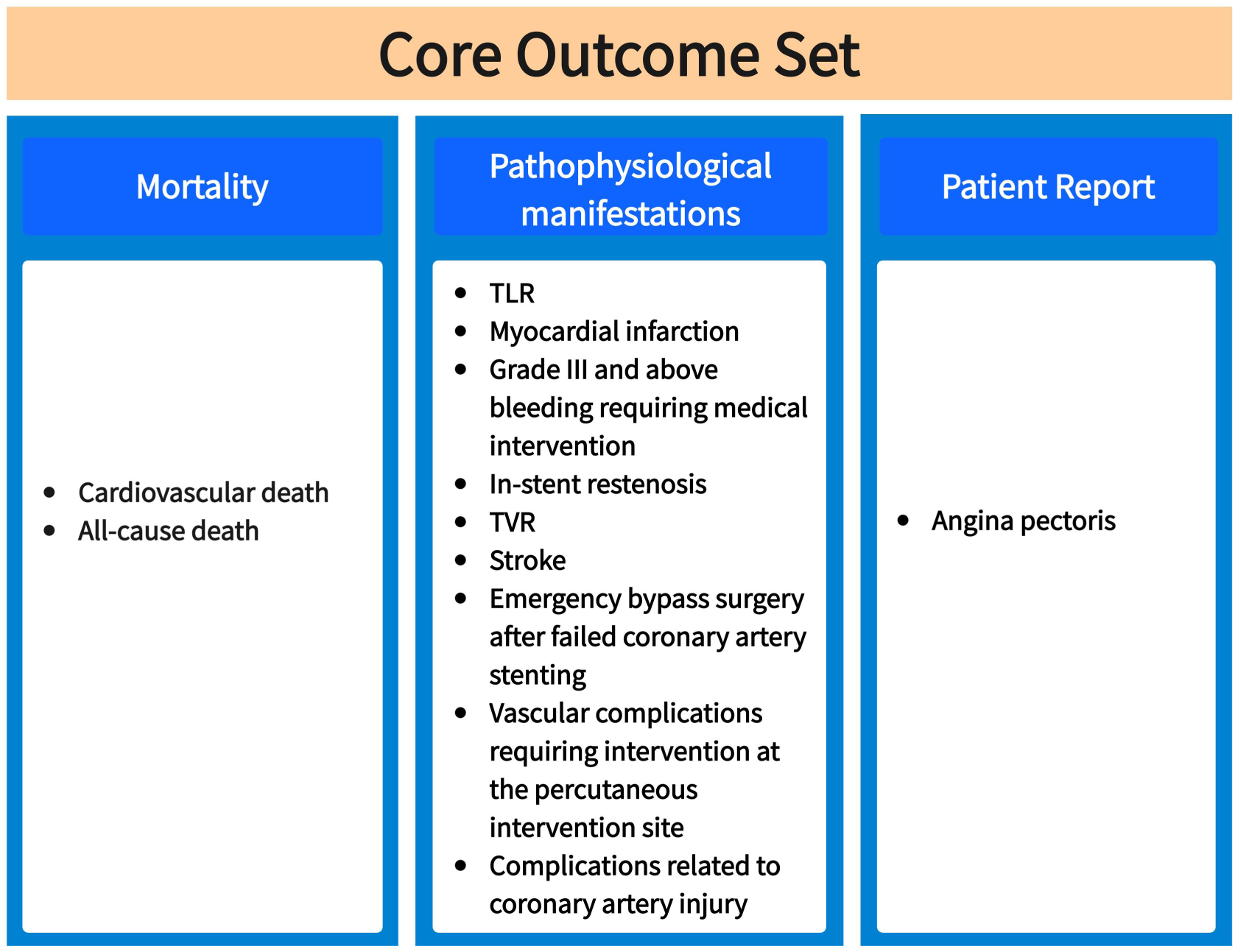
Final COS for coronary stent implantation for CHD.

## 4 DISCUSS

We established the first evidence-based, multidisciplinary panel of experts and patient-involved core outcome set for CHD stent implantation trials by integrating a systematic review of published literature with a mixed-consensus approach. This set of twelve core outcomes, consisting of four effectiveness and eight safety outcomes, will facilitate the standardization reporting of clinical trials and enable meta-analysis.

Among effectiveness outcomes, TLR, TVR, in-stent restenosis, and angina pectorisare are viewed as critically important by medical professionals and patients through the Delphi process. These results are consistent with previous research^[28,29]^ in which these outcomes were perceived as benefit outcomes that affect prognosis. Conversely, angina pectoris, a patient-reported outcome, was rarely reported in the previous studies.^[30,31]^

Among safety outcomes, all-cause death, cardiovascular death, myocardial infarction, stent thrombosis, and Grade III and above bleeding requiring medical intervention are the top five outcomes with the highest level of consensus through the Delphi process, which were preferred to report in most trials contributed to they were perceived as severe adverse outcomes affecting quality of life.

Several outcomes were excluded because of the disparate perception of their importance among professionals and patients. For instance, compared to the outcome of vascular complications requiring intervention at the percutaneous intervention site, acute coronary artery occlusion was voted out of the COS through consensus meeting. Given that the patients who participated in the consensus meeting also had a history of stent implantation, it is possible that its low ratings were attributable to patients who perceived the importance of the pain experience of arterial puncture compared with the adverse events of low incidence. Another example is the change in the importance of the outcome itself. In-stent restenosis was more critical than stent thrombosis to all the participants. It is possible that the application of drug-eluting or bioabsorbable stent, the new generation of stent, has significantly reduced stent thrombosis, which was considered to be a criticism of metal stents.^[32-34]^ This may be similar to those outcomes whose importance would change along with the progress of stent technology. Therefore, this core set will be updated as new evidence emerges.

The quality of Delphi studies depends mainly on the composition of the stakeholder panel, underlining the importance of including all relevant stakeholder groups representing diverse disciplines and reflecting the target population.^[12,35]^ In addition to medical professionals, we invited industry-related participants to bring additional insight into regulatory requirements and best industry practices. Clinical decisions should be based on the outcomes measured in clinical trials and those relevant to patients.^[36]^ However, patient representatives are seldom consulted both in Delphi and consensus meetings to determine the type of outcomes. We extensively involved patient representatives in every step of the project, ensuring the integration of patients’ views, e.g., by adding well-represented outcomes for patients. This way, the final COS is relevant to all stakeholders related to coronary stent implantation, and subsequent research supports the view of patients, healthcare professionals, and researchers.

This study has several strengths. Firstly, we proposed to set the inclusion criteria of optimal representatives and ultimately invited seventeen well-informed representatives to achieve greater response consistency, ensuring the reliability and authority of establishing the COS. Secondly, this study shows a high participation rate in the first round (100%), followed by 92% in the second round, although the method used is time-consuming and requires active participation. In order to achieve accuracy, a response rate of at least 70% is needed.^[37]^ Based on previously mentioned COS development studies in other diseases, response rates above 60% could be expected. ^[38-41]^ This response rate is a significant strength of our study.

There are some limitations. First, all the Delphi panelists were Chinese-based because we initially wanted to develop a COS of stent implantation intervention for China. However, many aspects of clinical decision-making in stent implantation for CHD are generalizable to other countries and health systems, and we reference relevant international standardized endpoint definitions and consensus documents. Second, this study generated a list of potential core outcomes through a literature review of systematic reviews, differing from the previous studies, which directly extracted outcomes from clinical trials. ^[42-44]^ This may cause the possibility of the omission of the original outcomes. However, we supplemented the original list of outcomes by conducting additional searches in the COMET database and consulting relevant guidelines/expert consensus. The present study results showed that only four additional outcomes were added after the first round, indicating that the potential core outcomes in the list before the Delphi survey achieved an extent saturation level.

## 5 Conclusions

This study has developed the first COS in coronary stent implantation intended for use in RCTs, systematic reviews, and clinical practice guidelines evaluating stent implantation for patients with CHD. This COS will facilitate the standardization of coronary stent implantation assessment to reduce heterogeneity in assessment in future research. Finally, this COS should be updated as new evidence emerges in this rapidly evolving field. Following the future research on standardized measurement of this COS, the use and evaluation of this COS should be carried out in the next step.

## Author Contributions

Xingli Wan and Liang Du co-chaired the process and conceived the consensus project. Xingli Wan and Liang Du designed the questionnaires and coordinated the Delphi and consensus meeting project. Xingli Wan drafted the manuscript. Zhengchi Li and Liang Du revised the manuscript. Xinyi Wang and Xia Li carried out the literature review. Youlin Long participated in the selection of outcomes from this stakeholder group. All authors have read and agreed to the published version of the manuscript.

## Acknowledgments

We are grateful to the cardiovascular medicine/surgery experts, journal editors, methodologists, industry-related experts, and patients with the living experience of coronary stent implant treatment who participated in this consensus project. Their gracious time and contribution are highly acknowledged. We would like to acknowledge Zhonglan Chen and Juan Du, who helped contact patients. We also thank all the participants who made this study possible.

## Data Availability

Additional data can be obtained from the supplementary materials. The deidentified participant data will not be shared.

